# Preliminary Development of a Multidomain Behavioral Intervention for Primary Prevention of Dementia in Midlife

**DOI:** 10.1101/2024.08.09.24311754

**Authors:** L.E. Korthauer, R.K. Rosen, I. Arias, G. Tremont, J.D. Davis

## Abstract

**Background:** An estimated 40% of global cases of Alzheimer’s disease and related dementias (ADRD) may be prevented through modification of 12 risk factors. Midlife risk reduction is particularly important given the long prodromal period for ADRD. However, population adherence to these health behaviors is low, and large, multidomain behavioral interventions have suffered from poor adherence. The goal of this study was to use qualitative methods to develop a personalized health education intervention (TEACH: Tailored Education for Aging and Cognitive Health), which is grounded in the Health Belief Model.

**Method:** We conducted six focus groups in which we presented health information about a fictitious person, including images conveying ADRD risk and descriptions of personal health belief factors. Participants provided feedback about the images, their understanding of ADRD risk, and interpretation of the health belief concepts. Results were summarized using a framework matrix.

**Results:** Groups found the presented risk information to be understandable and relevant to health behaviors. They had strong reactions to presented images and descriptions, and they provided alternative language and suggestions to improve clarity. Participants noted some overlap between health belief constructs but were able to identify connections between health beliefs and personal behaviors.

**Conclusions:** Results guided the TEACH intervention development project, which includes creating an explanatory framework for disclosing individuals’ ADRD risk factors and health belief profile. The long-term goal is to develop a multi-domain intervention to promote sustained health behavior change for primary prevention of ADRD.

## Introduction

The 2020 report of the Lancet Commission on dementia prevention stated that 40% of dementia cases globally may be attributed to 12 modifiable risk factors. These include depressed mood, diabetes, early life education, excessive alcohol consumption, hearing impairment, hypertension, obesity, physical inactivity, smoking, social isolation, exposure to toxins (including air pollution), and traumatic brain injury (Livingston et al., 2020). Correctly timed interventions to modify these risk factors could substantially reduce the global burden of Alzheimer’s disease and related dementias (ADRD), which are expected to affect 13.8 million people in the United States alone by 2060 (Association, 2023). Given that ADRD and related neuropathologies begin to develop 10-15 years or more before the onset of cognitive symptoms (Jack et al., 2013), risk factor modification in midlife and early late life is critical to dementia prevention.

Despite a robust body of observational research examining the effects of health behaviors such as diet, sleep, and physical activity on dementia risk, there are few randomized controlled trials for primary prevention of dementia (see Sanborn et al., 2024, for review). One of the best-known large, multi-domain interventions is the Finnish Geriatric Intervention Study to Prevent Cognitive Impairment and Disability (FINGER) trial, which included diet, physical activity, cognitive activity, and vascular disease management compared to healthy living education in 1,260 older adults who were cognitively unimpaired or had mild cognitive impairment (MCI). After two years, participants in the multi-domain intervention had 25% larger improvement on cognitive composite measures than controls (Ngandu et al., 2015). Replication studies are ongoing in several countries, including the United States (U.S.-POINTER study) (Baker et al., 2023), though efficacy data are not yet available.

While multi-domain interventions may increase target behaviors and reduce risk of dementia, adherence remains problematic. In the FINGER trial, the active intervention increased the frequency of target behaviors and, consequently, cognitive performance. However, only 19% of participants adhered to all components of the FINGER intervention (defined as 66% attendance of sessions) (Coley et al., 2019), and those who were not adherent to the intervention had poorer cognitive outcomes (Ngandu et al., 2022). Poor adherence poses a particular problem for dementia prevention, because health behavior change must likely be sustained for years or even decades to have a meaningful impact on dementia risk.

Grounding behavioral interventions in theories of health behavior change is one potential way to increase long-term adoption of brain healthy behaviors. One of the most widely used models of behavior change is the Health Belief Model (Rosenstock, 1974). This model identifies health beliefs including perceived threat of disease, perceived benefits of change, perceived barriers to change, and self-efficacy as mediators of health behavior change. Despite its widespread use in other areas of public health (Jones et al., 2014), the Health Belief Model has only rarely been applied to primary prevention of dementia. In a registry of U.S. adults aged 45+ who were interested in dementia prevention research, health beliefs including perceived future time remaining in one’s life, self-efficacy, ability to defer gratification, and consideration of future consequences were associated with lower ADRD risk and engagement in brain healthy behaviors (Zakrzewski et al., 2024). This is consistent with data from an Australian sample showing that self-efficacy and perceived susceptibility of disease mediated the relationship between dementia knowledge and behavior change (Bartlett et al., 2023). Educating participants about these health belief factors and associations between health beliefs and behavior may be a powerful inducement to behavior change.

The goal of this NIH Stage I behavioral intervention development study was to use qualitative methods to develop a theoretically-grounded, personalized multidomain health behavior intervention (TEACH: Tailored Education for Aging and Cognitive Health) targeted to middle-aged adults. We identified several health belief constructs from the Science of Behavior Change Research Network (Nielsen et al., 2018), which were used to create a personalized health belief profile that will be used as part of the TEACH intervention (Figure 1). In this study, we conducted focus groups to develop an effective framework for communicating this health information to adults in a way that was understandable and applicable to their health.

**Figure 1.**
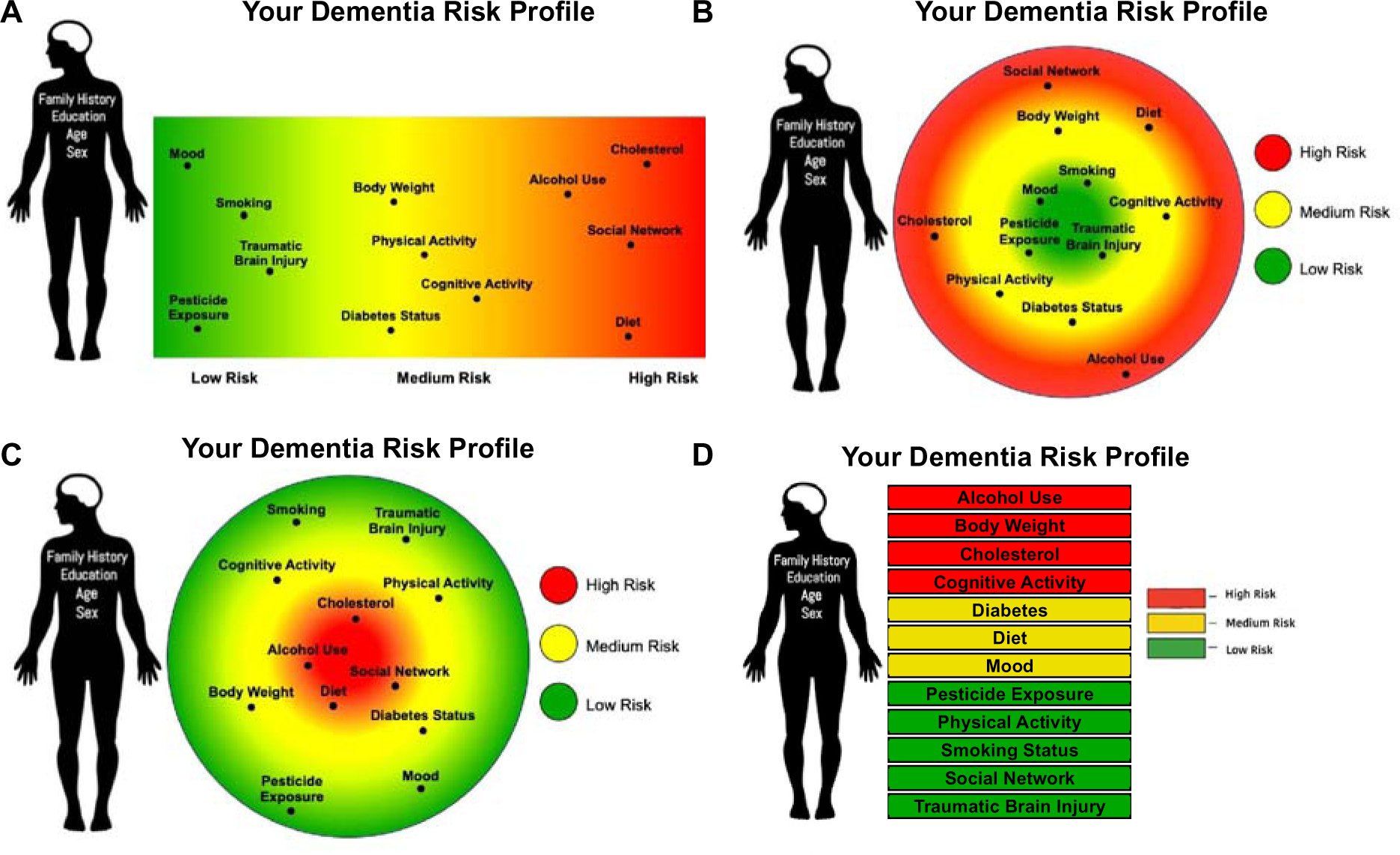
Images conveying a fictitious person’s risk of Alzheimer’s disease and related dementias, including non-modifiable and modifiable risk factors.

## Methods

### Participants

Participants were recruited from the Rhode Island Alzheimer’s Disease Prevention Registry through Rhode Island Hospital and from the surrounding Rhode Island community. The participants satisfied the following inclusion criteria: a) being between the ages of 45-69 years old; b) being cognitively unimpaired (Minnesota Cognitive Acuity Scale > 52); and c) being fluent in English. All study procedures were approved by the Rhode Island Hospital institutional review board.

### Focus Groups

Six focus groups consisting of 3-6 participants were held in person at Rhode Island Hospital or community locations. Each focus group was approximately 90 minutes long. Focus groups followed a standard discussion guide, including probes to explore and seek clarification. Each focus group was attended by two study investigators; one served as a facilitator to provide an overview of the group discussion and present questions and follow-up probes. A research assistant was present to record field notes and non-verbal communication.

During each focus group, participants were presented via PowerPoint slides with health information about a fictitious, gender-neutral person (“Taylor”). This included two primary domains of health information: 1) Taylor’s personal risk for ADRD and 2) their personal health beliefs. Regarding personal risk for ADRD, participants were shown four images meant to convey Taylor’s degree of ADRD risk for 12 modifiable (e.g., diet, physical activity, social network) and four non-modifiable (e.g., age, sex, family history) risk factors (Figure 1). Structured questions assessed participants’ understanding of the risk images, perception of which risk factors Taylor most needed to change, and opinions about the images used to convey the risk information.

Regarding Taylor’s personal health beliefs, participants were shown descriptions of seven health belief constructs: future time perspective, delay discounting, deferment of gratification, consideration of future consequences, response inhibition, executive control, and self-efficacy (Table 1). Each of these constructs corresponds to a measure from the Science of Behavior Change Research Network (Nielsen et al., 2018).

**Table 1.** Descriptions of health belief constructs.

| **HBM Domain** | **Construct** | **Layperson-Friendly Description** |
| --- | --- | --- |
| **Perceived threat** | Future time perspective | Future time perspective refers to how much time you believe you have left in life and how you would like to spend that time. |
|  | ADRD risk | N/A (conveyed by ADRD risk images) |
| **Perceived benefits and barriers** | Response inhibition | Response inhibition is your brain's ability to manage impulses. |
|  | Delay discounting | Delay discounting measures how much you want something right now, even if you could hold off for something better later on. |
|  | Executive control | Executive control is how well your brain can focus on a goal while ignoring things that could distract you from your goal. |
|  | Deferment of gratification | Deferment of gratification is your ability to put off something you want now in order to wait for something even better in the future. |
|  | Consideration of future consequences | Consideration of future consequences refers to how you think about the long-term consequences of your actions. |
| **Self-efficacy** | Self-efficacy | Self-efficacy is your belief in your own ability to reach a goal. |

*Note.* HBM: Health Belief Model. ADRD: Alzheimer’s disease and related dementias.

Each focus group was shown information about four of the seven constructs, with the order counterbalanced across focus groups so each health belief construct was discussed at least three times. A layperson-friendly definition (8^th^ grade reading level or below) was presented for each construct. Participants were asked structured questions to assess their understanding of the definitions, elicit alternative words or phrases to describe the constructs, draw links between the health belief constructs and Taylor’s health behaviors, assess the perceived relevance of the construct in their own lives, and examine perceptions of the constructs as fixed versus changeable. They were also shown three different images to convey Taylor’s level of each health belief construct (Figure 2). Structured questions assessed their understanding of the images and whether the images facilitated understanding of the written definitions of each construct.

**Figure 2.**
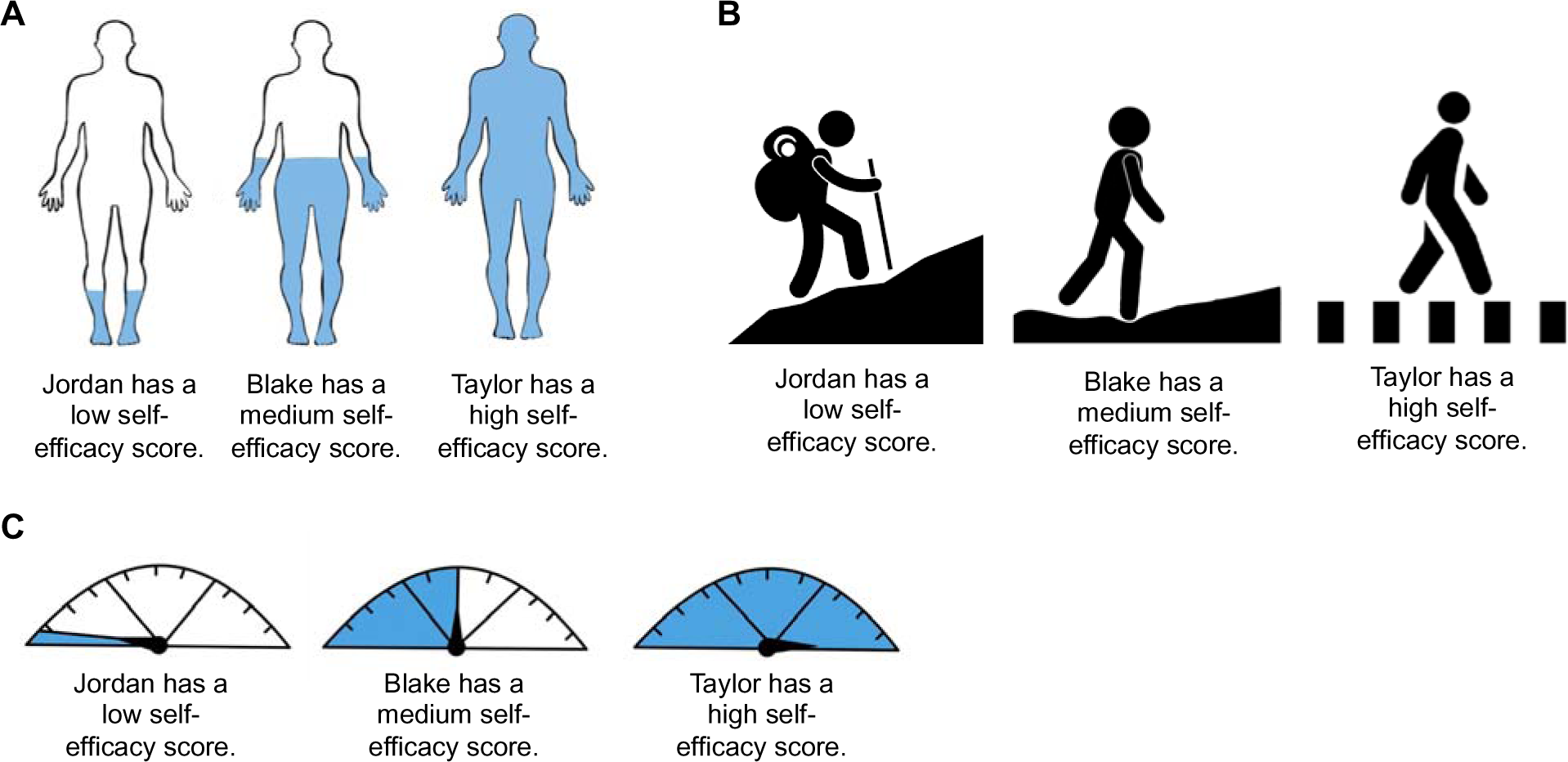
Images conveying fictitious individuals’ levels of health belief traits.

### Qualitative Analysis

Each focus group session was digitally recorded and professionally transcribed. All identifiers were scrubbed from the transcripts. Transcripts and field notes were reviewed by two members of the research team who are experienced in conducting focus groups and working with qualitative data. They reviewed data independently. Data were charted into a framework matrix for analysis (Gale et al., 2013; Rosen et al., 2023). One rater charted the data into a matrix by summarizing participant comments to each of the major questions posed in the focus groups. Responses were reviewed by the entire research team. Major trends were identified to determine clarity of health information provided, alternative ways of displaying and summarizing information, and preferences for explanatory images.

## Results

Demographic characteristics for the six focus groups are as follows: *M* age = 62, range = 47-69 years; 73% women; 96% non-Hispanic White; 42% from an ADRD prevention registry and 58% from the RI community. All participants had at least a high school education, and 35% had obtained a bachelor’s degree or higher.

Overall, focus groups demonstrated the understandability and relevance of information about ADRD risk and health belief constructs to non-expert participants. Participants had strong opinions and reactions to presented information. They were able to describe links between health belief factors and behaviors such as dietary choices or physical activity. Participants noted some areas of overlap between the health belief constructs, though subtle distinctions between the concepts were observed. Many expressed a desire to learn their own personal health information after seeing the information provided about a fictitious person.

### ADRD Risk Factors

Most participants were aware that some aspects of ADRD risk are modifiable. Diet and alcohol use were the most cited factors that were previously known. Participants were less aware of social network, history of traumatic brain injury, and pesticide exposure as modifiable ADRD risk factors.

Some participants expressed that listing education as a non-modifiable risk factor was confusing, because people can seek additional education throughout the lifespan. For example, this participant contrasts education level (8^th^ grade vs. college) with a person’s level of relevant health knowledge:

> “*The only one that surprised me was education. I wasn’t sure how that would actually fit into the picture. And I mean…if someone with an eighth-grade education would do worse than someone with a college education? Are we talking about that? Or is it education about health that you want to convey to people?” [participant #197; all alphanumeric identifiers are dummy codes so they cannot be linked back to actual study identifiers]*

### ADRD Risk Images

Participants were shown four different ways of indicating “Taylor’s” dementia risk. The first image showed a color bar with risk factors arranged from low risk (green) to high risk (red) (Figure 1A). Most participants understood the color bar image and were able to correctly identify what risk factors the person needed to work on.

> “*He has a lot of things that he could change if he had the ability to change…his cholesterol, alcohol use, social network, and diet…physical activity, too, it affects his body weight. A lot of those high things are under his control.”* [participant 155]

Participants were generally able to distinguish between the non-modifiable risk factors and modifiable risk factors.

> “*Well, you have to—at a point, you can’t change family history, age, or sex, but many of the things within the colored rectangle, they are modifiable.*“ [participant 113]

However, some participants found the rectangular color bar to be confusing. One participant wondered whether the horizontal axis indicated degree of risk while the vertical axis indicated the amount of each risk factor that Taylor had. A few participants found the words placed on the black silhouette of a person’s body to be confusing or alarming (”*Change it to a color. I like color. Black just seems so—I don’t know—negative*“ [participant 194] and “*The first body looks a little scary -- ‘cause it’s Halloween-ish.”* [participant 155]), but this was not a majority opinion. Some participants also wanted more information on the image about how the risk factors were operationally defined as low, medium, or high risk.

The next two risk images showed the person’s degree of risk arranged in concentric circles like an archery target (Figure 1B and 1C). The same information was presented with two color schemes, first with the low risk (green) items in the center of the target and second with the higher risk (red) items in the center. There was some disagreement about which color scheme was preferable, with most preferring higher risk (red) factors to be presented in the center of the bullseye.

> [In response to image 2; Figure 1B] *“Not to be fresh, but it looks totally opposite as to what you’re looking at. I mean, again, when we see circles like that, I think bullseye, so the red should be in the middle.”* [participant 161]

One participant concluded that the color scheme might depend on what information the researchers most want people to focus on:

> “*On this target type of visual, my eyes go to the center. Whether it was green or red, so you kinda want it to be the red, unless you want it to be the green. You know, unless you want them to see what they’re doing positively. It depends on…which side of the coin you want to look at.*“ [participant 183]

The final image was a vertical list of modifiable risk factors presented next to the black silhouette of a body, with risk factors arranged by high risk (red), medium risk (yellow), and low risk (green) (Figure 1D). Most participants found this image to be clear and understandable:

> “*This is cut and dry…but it’s nice at the same time.*“ [participant 127]

However, some participants disliked the lack of gradient in the degree of risk when it was categorized as low, medium, or high.

> “*The horizontal linear one, it just seemed to give you levels as well, where this -- you don’t know where the range is high is only high. There’s no variation within the high*.” [participant 161]

When asked to vote on their preferred image, 12 participants voted for image #4, seven for image #1, and two for image #2.

### Health Belief Constructs

Participants expressed generally strong understanding of each of the seven health belief constructs. They were able to articulate differences between the concepts, though several were conceptually connected (i.e., consideration of future consequences and deferment of gratification; response inhibition and delay discounting), and some participants perceived them as overlapping or duplicative. For some constructs, particularly future time perspective, they had mixed opinions about whether having more of the trait would make a person more or less likely to engage in a health behavior. They also indicated that there is a situational component to several of these constructs, wherein a person may have a high level of a particular trait but may not always optimally use that trait when making day-to-day decisions about health behavior changes. For example:

*“You’re human and you’ve got your impulses, and whichever your impulses are, it’s hard to do it sometimes. But you have to. And then you fight with your mind….And you’re like…I won’t. Nope, nope. No. Yeah, yeah…and then you [have to] do it [again] the next day.”* [participant 163]

Participants also provided alternative language to ensure descriptions were understandable. When asked which construct is most relevant to their own health, each of the seven concepts was considered primary by at least one participant. The most cited constructs were consideration of future consequences, executive control, and response inhibition.

### Generalized self-efficacy

The concept of self-efficacy was generally well understood. Many participants suggested the alternative term “self-confidence” as more widely known, though several participants recognized subtle distinctions between self-efficacy and self-confidence.

> “*Some people have a lot of confidence in themselves, but…are really not competent.”* [participant 179] *“You make a really interesting point there. You can have self-efficacy without having confidence, and vice versa. Self-efficacy isn’t really your belief in your ability to reach a goal. It’s your ability to reach a goal.”* [participant 139]

Most participants expressed that self-efficacy is something that is under one’s control and noted that higher self-efficacy scores are likely to translate to greater engagement in healthy behaviors.

### Deferment of gratification

Participants found the concept of deferment of gratification to be intuitive. They were able to relate a high, medium, or low score to how a person may or may not engage in health behaviors. Many provided examples about dietary choices as well as physical activity and alcohol consumption. Some participants noted that a person’s ability to defer gratification may fluctuate based on situational factors. Opinions were mixed as to whether deferment of gratification is a fixed trait or malleable. Some people felt that it could be controlled, and others discussed circumstances in which it might not be, such as if a person has a neurodevelopmental disorder such as Attention Deficit/Hyperactivity Disorder.

> “*Listening to you say that makes me think of those people who have, for example, ADHD…you know, they’re just wired to basically have instant gratification and not deferment. And that’s just not within their control*.” [participant 182]

### Consideration of Future Consequences

Participants demonstrated a good understanding of the concept of consideration of future consequences. Several identified parallels with deferment of gratification, though subtle differences between the constructs were noted.

> “*That word ‘defer,’ to me, makes that a little different…this [consideration of future consequences] is less of an active term to me. Whereas deferment is more of an active type of term. So, I do think that this one is something that you could change because…you’re up in your brain thinking about things all the time. You could consider all sorts of things all the time.”* [participant 113]

Participants also identified ways in which consideration of future consequences would affect health behaviors.

> [If a person has a low score] “*They would be less likely to follow up and do things to improve our health…they might just ignore whatever their doctor tells them or what other health professionals…’you’re going to die anyway’…I’ve heard that from my relatives. ‘Nobody lives forever, you know. Might as well enjoy myself.*’” [participant 194]

### Response inhibition

Participants expressed understanding of the concept of response inhibition, though several noted some overlap with other constructs, including deferment of gratification and consideration of future consequences. One participant noted that a high score on something with “inhibition” in its name might be hard for people to understand. Another participant suggested the phrasing “suppressing negative impulses” might be more understandable.

Participants had mixed opinions about whether response inhibition is a malleable trait. Some people expressed that response inhibition is something that one’s brain simply does and is outside of one’s direct control. Others partially agreed with that position but said that a person can be taught strategies to more effectively use response inhibition in certain situations:

> *“Everybody’s got impulses, positive and negative…but it’s the ability to suppress those impulses, you know?…You have to get yourself out of that environment that’s going to get you into trouble.”* [participant 124]

### Delay Discounting

The concept of delay discounting was confusing to some, though not all, participants. Several said that the definition was more ambiguous than the other health belief constructs. The word “discounting” was perceived as confusing by several participants. The interpretation of a high versus low score was also confusing to some:

> *“So delayed discounting shows how much you want something right now, even if you could hold off for something better later on. So are you saying he had a high score of wanting something right now? Or he had a high score of holding off for something better later on?”* [participant 196]

Alternative language, such as “self control” was suggested. Participants noted overlap between this construct and response inhibition and delayed gratification.

### Executive Control

The concept of executive control was well understood by most participants. Again, some similarity was noted between this construct and other health beliefs such as response inhibition. However, some conceptual differences between these were noted:

> “*Executive control…isn’t really the same as self-control, because executive control is more how your overall brain works and how it executes things…I think response inhibition and executive control are connected in some ways…think of response inhibition as being more of a short-term thing, with executive control as more long-term*.” [participant 179]
>
> “*I think of executive control as getting yourself to do and to complete things, whereas response inhibition is getting yourself to not do things*.” [participant 139]

Executive control abilities were considered an important factor in engagement in health behaviors by most participants. Participants differed in the degree to which they believed a person could modify their executive control abilities.

### Future Time Perspective

Comments suggested that there were a variety of responses to this concept. Some participants felt that a high score on future time perspective (i.e., feeling like there is a long amount of time remaining in one’s life) would motivate a person to make health behavior changes. Others felt the opposite, that feeling that there is a large amount of time remaining would decrease motivation to make health behavior changes now.

> “*See, if I think I have all the time in the world, I don’t feel particularly motivated to have to do anything, ‘cause I always have an opportunity down the road to do it*.” [participant 179]
>
> “*If I said, ‘Oh, my God. I only have six more months to live,’ I’m eating everything. I’m going – that’s it…If I said, ‘Oh, gosh, I’m gonna be around for 30 more years,’ I better start working on this now so that I have, you know, 29 really good years…to make sure I make it to the 30*.” [participant 180]

### Images to Convey Level of Each Health Belief Construct

Participants were also shown three sets of images to convey the level of each trait a fictitious person has. The first set of images showed silhouettes of a person with a low, medium, or high level of blue color (Figure 2A). The second set of images showed a person walking on flat ground (intended to indicate a high amount of a trait that would facilitate health behavior change), a slight hill, or up a mountain (indicating a low amount of a trait that would facilitate health behavior change) (Figure 2B). The third set of images showed a speedometer indicating low, medium, or high amounts of a health belief trait (Figure 2C).

The first and third sets of images (silhouettes and speedometers, Figures 1A and 1C) were understood by most participants. The second set of images was less well understood, with greater disagreement among participants about how to interpret the images and connect them to levels of health belief traits. Some participants inferred that the hiker on the mountain had traits that would pose barriers to health behavior change: “*Jordan over there is struggling. Jordan knows he’s got struggles and it’s…gonna be a mountain just to get through life.*” [participant 127]. Others viewed the hiker as someone who would be more likely to achieve their goals: “*You think of people who are hiking as people who are, you know, determined and they have goals, and they set them, and they’re working hard*.” [participant 196]

Multiple individuals expressed that no images were needed to understand the health belief constructs and that the images distracted from the concepts themselves. Some participants expressed very negative reactions to the images. For example, reactions to the first set of images (Figure 1A) included, “*It’s a bad graphic.*” [participant 196], “*They’re kinda creepy.*” [participant 180], and *“I see blue socks*.” [participant 183]. Reactions to the third set of images (Figure 1C) were similarly varied. Some participants commenting that the speedometer removed the human element of the health belief traits: “*[You] dehumanized it, though…where’d Taylor go?*” [participant 161] and “*It’s like a sports car, but we’re talking about a person here.”* [participant 140]. However, others found this image the most intuitive to understand: *“If we’re going to go with a graphic, I like this one better because it’s not as distracting with people, like thinking about the people. This one is a…gauge.”* [participant 183] and “*That was better than the bodies [from images 1 and 2; Figure 2A and 2B]*” [participant 155].

When asked to vote on a preferred set of images, participants were relatively evenly split, with nine preferring the first set of images (silhouettes), two preferring the second set (hiker), and eight preferring the third set (speedometer).

### Considerations for Individual Score Disclosure

Most participants expressed interest in learning their personal scores on the ADRD risk assessment and health belief constructs. One person expressed that learning their personal information would make them feel “empowered.” Multiple participants expressed that they would want emotional support and actionable strategies to manage their ADRD risk. Some participants reflected that their response to a hypothetical disclosure would depend on their scores.

> *“It would be great if I scored high on everything. But if all of my scores were low I’d be like, ‘Ugh. Again.’ You know? So it depends on how it’s presented…I would need psychological and emotional support, and a big support team if I was told. Especially if you’re already somebody who is not well and not healthy…Not just, ‘you need to do this better.’ [but] how?”* [participant 196]
>
> “*I’m assuming if someone’s giving you these results, they’re saying to you, ‘We know the risk factors, but we also know that…if we improve these, that your chances lessen.’…So this is our starting point. Let’s be positive. It’s our starting point, but we can move forward and improve those…if I was all red [high risk], I’d probably go home and cry, but then I’d know I have somewhere to go. So I’m at the bottom, but I can go up maybe, if I was all red*.” [participant 127]

## Discussion

These results represent the first step of a Stage I intervention development study for TEACH, an intervention being designed to educate people about modifiable risk factors for ADRD and health beliefs that may affect engagement in relevant health behaviors. Using focus groups, we examined participants’ understanding and reactions to the health information of a fictitious character. This included disclosure of modifiable and non-modifiable dementia risk factors as well as health belief concepts that may impact engagement in modifiable health behaviors.

Focus groups demonstrated good understanding of ADRD risk information and found it to be relevant to health behavior which is encouraging as perception of risk is critical for making individual health behavior change per the Health Belief Model (Rosenstock, 1974). This is consistent with prior research examining health education in the context of ADRD risk, though past work has primarily focused on disclosure of biological risk factors such as *APOE* ε4 or AD biomarker status. For example, the REVEAL study showed that 63% of participants recalled their *APOE* ε4 status after one year (Eckert et al., 2006), and those who were at higher genetic risk were significantly more likely to self-report health behavior change (Chao et al., 2008). Other studies have found minimal or absent health behavior change after ADRD risk disclosure (Popescu et al., 2024; Sng et al., 2019), suggesting that knowledge of ADRD risk alone is insufficient to motivate behavior change.

Focus group participants were generally able to draw connections between personal health belief factors and health behaviors. Most individuals were able to generate specific examples linking a high or low score for a given health belief factor to a hypothetical health behavior. Participants described overlap between several of the constructs. For example, consideration of future consequences and deferment of gratification were perceived to similar, as were response inhibition and delay discounting. Several participants described subtle distinctions between these concepts, which will be used to develop our explanatory framework for educating individuals about these constructs.

One of the health belief factors that generated the most varied responses was future time perspective. Some participants expressed that a higher future time perspective (i.e., more perceived future time remaining) would be a motivator for health behavior change, while others said that this would reduce motivation. Although a recent meta-analysis reported small-to-medium effect sizes between future time perspective and health behavior outcomes, cultural variability was a significant moderator of these effects (Andre et al., 2018). Thus, individual-level associations between future time perspective and engagement in health behaviors may be highly variable and partially dependent on age, gender, and cultural background. Importantly, this does not preclude us from including future time perspective in this type of behavioral intervention. The goal is to build individuals’ awareness of health belief traits and their relationships to health behaviors, regardless of whether an individual perceives a given trait as a barrier or facilitator of behavior change.

Using the feedback provided by the focus groups, we have created an explanatory framework for disclosing individuals’ personal risk for ADRD and educating them about their personal health beliefs. Based on the preferences of most respondents, we will use image #4 (the vertical list of risk factors in red, yellow, and green) to convey participants’ individual ADRD risk. Some participants expressed a preference to convey the magnitude of risk conferred by each risk factor. However, the study team agreed that this may be unrealistic and misleading in practice, as relative risk conferred by each factor interacts with person-level characteristics such as age, sex, socioeconomic status, and medical history. Thus, we will categorize risk factors into low, medium, and high risk and alphabetize the list of risk factors within each category. We have also clarified language used when conveying ADRD risk and health belief information based on participants’ feedback. Although several of the health belief constructs were perceived to be overlapping, they were understood by the majority of participants. Thus, we will retain all the measures in the explanatory framework and make decisions about whether to consolidate or eliminate any after the next phase of qualitative data collection and analysis.

The next phase of this study is to disclose individuals’ personal ADRD risk and health belief information to them using our explanatory framework. Individuals will complete a health belief assessment, and a licensed psychologist will disclose their ADRD risk and personal health belief information to them. We will then conduct individual interviews using a phenomenographic method, which focuses on the variations in how people learn and understand concepts. We will examine the acceptability, appropriateness, and applicability of the presented information to participants’ engagement in health behaviors important for ADRD prevention. This will be used to further refine the explanatory framework for our intervention, which focuses on educating people about their health beliefs and how to change their behavior to reduce risk for ADRD. We will conduct a randomized controlled trial to assess the feasibility of delivering this intervention versus basic health education alone on proximal outcome measures including perceived threat of ADRD, dementia knowledge, and self-efficacy.

The data from this study represents a first step in this Stage I intervention development project. One limitation of this study is the lack of racial and ethnic diversity in our sample. We are conducting a follow-up study with a more representative sample that additionally examines the association between social determinants of health, these health belief factors, and health behaviors. We will use the additional qualitative data from this follow-up study to further refine the explanatory framework and address structural barriers to health behavior change. Another limitation is that we presented fictitious data, rather than individuals’ personal health information. Reactions to personal health information may vary, and this is an important area for future research. Finally, although the health belief traits are presented as separate constructs, they are intercorrelated (Zakrzewski et al., 2024). Future research may consider more detailed behavioral phenotyping of these health belief constructs and their inter-relationships to guide intervention development.

In summary, we demonstrated that focus group participants understood images demonstrating a fictitious person’s degree of risk of ADRD based on 12 modifiable risk factors. They could also describe differences between health belief concepts and provided example language to make these concepts more accessible to a non-expert audience. They made connections between health belief traits and specific health behaviors. Overall, participants expressed enthusiasm and interest in learning about their own personal risk profiles to lower risk for ADRD and improve their lifestyles. This information will be used to develop an explanatory method for disclosing participants’ personal health information to them as part of the TEACH intervention.

## Data Availability

All data produced in the present study are available upon reasonable request to the authors

